# To what extent is the altitude at which we live associated with 10-year cardiovascular risk?

**DOI:** 10.1101/2021.04.22.21255947

**Authors:** Andrea Cevallos Guerrero, Heidi Angela Fernández, Ángela León-Cáceres, Luciana Armijos-Acurio, Carlos Erazo, Ruth Jimbo-Sotomayor, Hugo Pereira-Olmos, Henry Pineda-Abarca, Erika Quishpe-Narváez, Xavier Sánchez, Carmenza Sevilla, Betzabé Tello, Ana Torres-Castillo, Tatiana Villacrés, Iván Dueñas-Espín

## Abstract

**Introduction:** There is evidence that demonstrates lower incidence rates of cardiometabolic factors at the highlands. There are no studies which correlate the altitude with formally calculated cardiovascular risk by a meter-by-meter approach. Under the hypothesis that cardiovascular risk is inversely associated with altitude, this study was aimed to assess such association.

**Materials and methods:** Cross sectional study using data from the Ecuadorian National Health Survey of 2012. We analyzed available information of adults of ≥ 40 to 60 years old who have sociodemographic, anthropometric, cardiovascular risk factors, and laboratory biomarkers that were included in the survey. We assessed the independent association between altitude of the housing in which survey participants lived at, on a meter-by-meter approach, and cardiovascular health risk at ten years, formally calculated by Framingham equations.

**Results:** Linear regression model showed that participants had 0.0005 % less probability of developing cardiovascular disease at 10 years *per* each increase in a meter in the altitude that participants live at (p<0.001), adjusted for sex, age, ethnicity, educational level, availability of social security, immigrants in family, area, income quintile, overcrowding (≥ 7 inhabitants in the house), any alcohol consumption, history of hypertension, body mass index, hematocrit, and triglycerides.

**Conclusion:** From a public health perspective, altitude at which individuals live is an important health determinant of cardiovascular risk. Specifically, *per* each increase of 1000 m in the altitude that people live at, there is a reduction of almost half a percentual point in the cardiovascular risk at 10 years.

## Introduction

Cardiovascular diseases (CVDs) are the leading cause of death worldwide, claiming the lives of approximately 17.9 million people annually [1]. They account for roughly 50% of non-communicable disease deaths [2,3] and more than three quarters of these deaths take place in low- and middle-income countries [4]. CVDs include coronary heart disease and stroke, among the most important, all of which produce substantial health and economic burden globally [2].

It is widely accepted that CVDs are closely related to social determinants (SDs) of health, which are defined by the World Health Organization (WHO) as “circumstances in which people are born, grow, live, work, and age, and the systems put in place to deal with illnesses” [5–7]. Scientific evidence has shown that SDs such as socioeconomic status, demographic characteristics, education, work environment, health care services and housing may be associated with the development of CVDs; however they are not routinely acknowledged during risk assessment in clinical practice [8–10]. At the same time, a lot of efforts are directed to identify and treat individual-level risk factors [11]. In this regard, analyzing SDs is crucial for a complete cardiovascular clinical assessment.

Environmental determinants are key risk factors linked to CVDs. Although they are generally understudied and overlooked [10], growing evidence suggests that variations in factors such as temperature, altitude, climate and pollution, among others, remain significant determinants of cardiovascular risk [12,13]. Specifically, altitude continues to be a factor closely associated with cardiovascular risk. Several studies support an inverse association between altitude and cardiovascular risk factors [14–18], but the required time of exposure for a protective effect is unclear.

Overall, there are two scenarios, a rapid change to high altitude and a progressive adaptation to it. It is well known that patients with CVDs traveling from areas at sea level to the ones at high altitude –defined as the terrestrial elevation at which oxygen hemoglobin saturation decreases below 90%; corresponding to an altitude of about 2500 m– [19] have an increased mortality risk as a consequence of lower physiological reserves [20]. Several explanations have been proposed. Individuals with a reduced physiological reserve and subsequent difficulty to adapt to hypoxic conditions at high altitudes can present decreased functional capacity due to impaired pulmonary gas exchange and/or cardiac output. However, there is scarce epidemiological data discussing morbidity after traveling to altitude areas among this population [20].

Considering that about 400 million people live at an altitude of over 1500 meters above sea level and around 150 million people live in regions higher than 2500 m [21,22], altitude remains as an important health determinant that requires proper analysis. This population has managed to survive at high altitudes for generations; therefore, contrary to the above-mentioned statement, scientific evidence suggest that they must have developed significant adaptations to lower oxygen levels to survive [13,23].

Long term exposure to high altitudes seems to be associated with a lower incidence of obesity [14,17], high blood pressure [16,24], metabolic syndrome [15,18], diabetes [15], and, even, a better expectancy of life [25]. In fact, living at a geographically higher elevation was also associated with a lower risk of developing metabolic syndrome [18]. Specifically, no studies have focused on the association between formal cardiovascular risk estimation and altitude, and most of those studies are only focusing on the association between just a few categories of altitude, making it difficult to understand the association and its real magnitude on a meter-by-meter basis.

With this background we hypothesized that altitude is inversely related with cardiovascular risk. Using the databases of the Ecuadorian national health survey of 2012 we aimed to assess the association between altitude and cardiovascular risk among the Ecuadorian adult population, of ≥ 40 to 60 years old on a meter-by-meter basis.

## Materials and Methods

### Design

Cross sectional study using data from the Ecuadorian National Health Survey (NHS) of 2012.

### Population and databases

We included in our study available information of adults of ≥ 40 to 60 years old who have sociodemographic, anthropometric, cardiovascular risk factors, and laboratory biomarkers that were included in the in NHS in 2012 (see flowchart in the **S1 Figure**).

The Ecuadorian national health survey was a cross-sectional study conducted in 2012 which involved nationally representative samples of the civilian noninstitutionalized Ecuadorian population, the details regarding the survey could be found elsewhere [15,26]. The survey was conducted by the National Institute of Statistics and Census using a stratified, multistage cluster design [26].

### Measurements

We included in our analyses sex, age, ethnicity, educational level, availability of social security, immigrants in family, area, income quintile, overcrowding (≥ 7 inhabitants in the house), any alcohol consumption, history of hypertension, body mass index, hematocrit, and triglycerides. Ethnicity was self-reported and classified as Indigenous, Afroecuadorians, Montubios and Mestizos. Standardized blood pressures were obtained by sphygmomanometer, and mean blood pressures from the last 2 measurements were used in the Ecuadorian NHS. Blood lipids were measured enzymatically. Detailed specimen collection and processing instructions are described in the NHS [26]. Anthropometry was assessed in a standardized way, and body mass index (BMI; in kg/m^2^) was calculated.

The main explanatory variable was altitude on a meter-by-meter assessment, using information from the altitude of the housing in which survey participants lived [15,26]. Altitude was obtained by georeferencing, according to the methods that have been described elsewhere [26].

### Cardiovascular Health Metrics

Cardiovascular health metrics included blood pressure, blood glucose, and lipid profile. High blood pressure was defined with a self-reported history of hypertension, diabetes mellitus was defined when the fasting glucose was higher or equal to 126 mg/ dL [27]. Risk of CHD and stroke or CVD at 10 years were calculated by Framingham equations (available at http://cvrisk.mvm.ed.ac.uk/help.htm) [28,29].

### Sample considerations and statistical analyses

Using the prevalence of high blood pressure as the methodological variable, a random sample of 3601 individuals is sufficient to estimate, with a confidence of 95% and a precision of +/-1 percentage units, a population percentage that will foreseeably be around 9.3% [26]. In percentage of necessary replacements, it has been foreseen that it will be 10%. The final sample was 3661 participants included in the study assuring statistical power (see Figure 1 and the online supplementary material for further details) (see). The sample calculation was performed using the GRANMO population calculator http://www.imim.cat/ofertadeserveis/software-public/granmo/ [30].

**Figure 1.**
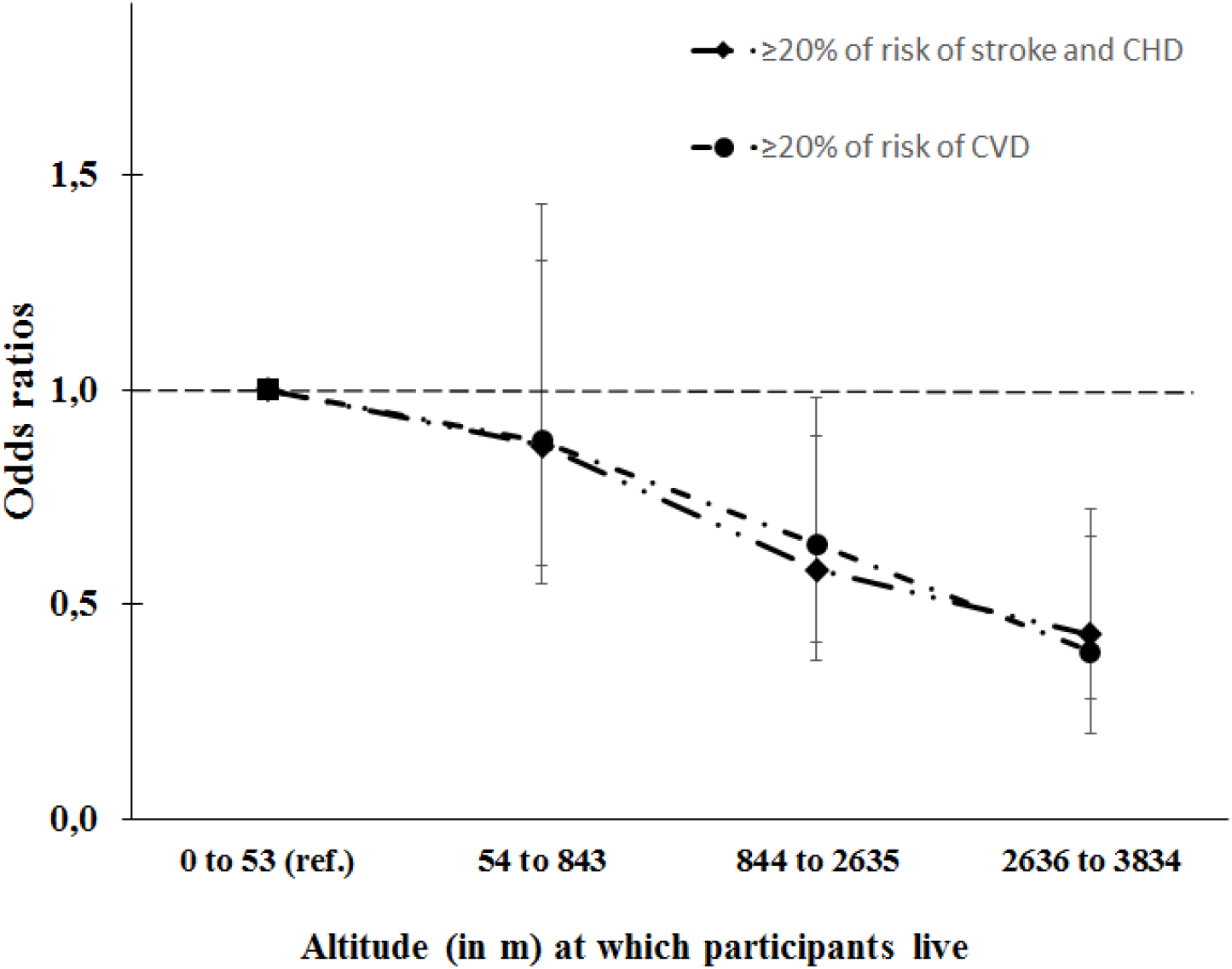
Adjusted odds ratios of high risk (≥ 20%) of stroke and/or CHD and high risk of CVD at 10 years by altitude categories, according to two multivariate logistic regression models, considering the reference an altitude of the residence of <54 m.

First, we tested the bivariate associations between altitude (both categorized four quartiles and as continuous variables) and the percentage of cardiovascular risk (CHD, Stroke and CVD) at 10 years, using analysis of variance and linear regression. Despite altitude and cardiovascular risk were non-normally distributed (**S1 and S2 Figures**), and prior to building multivariable models, we used generalized additive model analyses to test whether the shape of the association between altitude and cardiovascular risk was linear.

Then, we used multivariable linear regression to assess the independent effect of altitude on CVD. We tested a list of potential confounders, including age, sex, ethnicity, economic quintile status, social security availability, overcrowding (≥ 7 inhabitants *per* home), emigration, smoking and alcohol consumption, hypertension, hematocrit, and triglycerides. From these variables, we built a “saturated model”, which included: *(i)* variables independently related to the exposure or the outcome (p-value < 0.05), or that modified (>10% change in coefficient) the estimates for the remaining variables, and *(ii)* those variables that were considered important to all researchers. Then, we built a “parsimonious model” using a backward stepwise elimination process from the “saturated model” removing variables (one at a time) if Wald’s p-value <0.05. We tested both models, saturated versus parsimonious by the likelihood ratio test which was used to choose the final model and for the interpretation of the results. We tested goodness-of-fit of the final models.

We also built logistic regression models, including all significant variables that were identified in the final linear regression models, using the dichotomous outcome variable “high cardiovascular risk” when there was ≥20% risk of CHD and stroke or CVD at 10 years.

We performed several sensitivity analyses. Thus, we ran the final model stratifying by sex and by excluding *(i)* those who were living at less than 53 m (first quartile of altitude), *(ii)* those who were living at higher areas than 2635 m (fifth quartile of altitude), *(iii)* those with self-reported diagnosis of hypertension, and *(iv)* those with elevated LDL cholesterol (≥160 mg/dL) [31].

To identify potential effect modification, we stratified main analyses according to sex, history of hypertension, and smoking status. The analyses were performed using Stata 16.1 (Stata Statistical Software: Release 16.1; StataCorp LP, College Station, TX, USA).

The NHS which we used was performed in 2012. The specific ethical issues have been described elsewhere [26].

## Results

A total of 3661 patients were included. Most patients were women (69%), with a mean (SD) age of 45 (7) years, the majority were mestizo population (83%), with primary or lesser education (50%), only 35% had social security, mostly from urban areas (62%). The median (P25 to P75) altitude at which participants lived was 843 (53 to 2635) m. Regarding harmful health-related habits, most participants (90%) stated they have consumed alcohol at some point during their lifetime. There was also a tobacco smoking prevalence of 45%. The prevalence of self-reported history of hypertension and diabetes (as measured by fasting glucose ≥126 mg /dL) was 24% and 5%, respectively. Mean (SD) BMI was 28 (5) kg/m^2^, and mean total cholesterol was 196 (41) mg/dL (**S1 Table**). The prevalence of high (≥20%) cardiovascular risk (stroke or coronary heart disease) at 10 years was 7%.

In the bivariate analysis, increasing altitude was associated with lower median percentage cardiovascular risk at 10 years, as it was 8% in the lowest quartile of altitude (<54 m) and 6% in the higher quartile of altitude (>2635 m) (p-value<0.001), and with a lower median risk of stroke or coronary heart disease at 10 years, being 7% in the lowest quartile of altitude (<54 m) and 5% in the higher quartile of altitude (>2635 m) (p-value<0.001). Additionally, in the crude linear regression model we found a linear dose-response shape, with a reduction of −0.0005 percentual points in CVD risk *per* each increase in a meter in the altitude at which participants live, p-trend<0.001; interestingly, each cardiometabolic risk factor seems to improve with increasing altitude (**Table1 and Table 2**).

**Table 1.** Description of the sample across altitude categories.

| Variable | Altitude in m<br>(n = 3661) |  |  |  | p-value |
| --- | --- | --- | --- | --- | --- |
|  | 0 to 53<br>n=919 | 54 to 843<br>n=914 | 844 to 2635<br>n=914 | 2636 to 3834<br>n=914 |  |
| Female sex, n (%) | 604 (24) | 634 (25) | 647 (26) | 647 (26) | 0.061** |
| Age, mean (SD) | 46 (7) | 44 (8) | 45 (7) | 45 (7) | <0.001* |
| Ethnicity |  |  |  |  |  |
| Indigenous, n (%) | 11 (3) | 107(32) | 83 (25) | 136 (40) | <0.001** |
| Afroecuadorians, n (%) | 89 (56) | 47 (29) | 11 (7) | 13 (8) |  |
| Montubio, n (%) | 61 (50) | 54 (44) | 3 (2) | 4 (3) |  |
| Mestizo, n (%) | 758 (25) | 706 (23) | 817 (27) | 761 (25) |  |
| Education level |  |  |  |  |  |
| Higher than secondary education, n % | 198 (33) | 92 (15) | 159 (27) | 147 (25) | <0.001** |
| Secondary education, n % | 342 (27) | 307 (25) | 312 (25) | 283 (23) |  |
| Primary or lesser education, n % | 379 (21) | 515 (28) | 443 (24) | 484 (27) |  |
| Social security availability, n (%) | 318 (25) | 299 (24) | 347 (27) | 300 (24) | 0.063** |
| Any immigrant in the family (no immigrant is the ref.) | 8 (12) | 10 (15) | 22 (33) | 27 (40) | 0.001** |
| Rural area (urban is the ref) | 161 (12) | 442 (32) | 366 (26) | 423 (30) | <0.001** |
| Income quintiles |  |  |  |  |  |
| First quintile (the poorer group), n (%) | 146 (18) | 271 (34) | 175 (22) | 210 (26) | <0.001** |
| Second quintile, n (%) | 157 (22) | 241 (33) | 181(25) | 141(20) |  |
| Third quintile, n (%) | 209 (29) | 210 (29) | 168 (23) | 139 (19) |  |
| Fourth quintile, n (%) | 237(33) | 120 (17) | 179 (25) | 177 (25) |  |
| Fifth quintile (the richest), n (%) | 168 (24) | 70 (10) | 211(30) | 247(35) |  |
| ≥ 7 members in the family, n (%) | 130 (21) | 212 (35) | 136 (22) | 133 (22) | <0.001** |
| Harmful habits |  |  |  |  |  |
| Alcohol consumption, n (%) | 839 (26) | 799 (24) | 823 (25) | 806 (25) | 0.041** |
| Tobacco smoking, n (%) | 452 (28) | 407 (25) | 415 (25) | 369 (22) | <0.002** |
| Comorbidities |  |  |  |  |  |
| History of hypertension, n (%) | 675 (24) | 664 (24) | 712 (26) | 713 (26) | 0.014* |
| History of diabetes, n (%) | 848 (24) | 867 (25) | 880 (25) | 885 (25) | <0.001* |
| Anthropometry |  |  |  |  |  |
| Weight in Kg, mean (SD) | 69 (13) | 69 (13) | 68 (12) | 66 (12) | <0.001* |
| Height in cm, mean (SD) | 156 (9) | 156 (9) | 154 (8) | 154 (8) | 0.119* |
| BMI in Kg/m2, mean (SD) | 28.36 (5) | 28.29 (5) | 28.58 (4) | 27.90 (4) | <0.001* |
| ≥18.5 to <25, n (%) | 202 (26) | 204 (26) | 167 (22) | 199 (26) | 0.087 |
| <18, n (%) | 6 (35) | 3 (18) | 3 (18) | 5 (29) |  |
| 25 to <30, n (%) | 376 (25) | 346 (23) | 400 (26) | 409 (27) |  |
| ≥ 30, n (%) | 277 (26) | 258 (25) | 278 (27) | 237 (23) |  |
| Systolic pressure, mean (SD) | 125 (17) | 122 (16) | 121 (16) | 121 (15) | 0.001* |
| Diastolic pressure, mean (SD) | 77 (10) | 76 (11) | 75 (10) | 75 (10) | 0.057* |
| Laboratory Tests |  |  |  |  |  |
| Hematocrit in %, mean (SD) | 41 (4) | 41 (4) | 43 (4) | 45 (4) | 0.633* |
| Glucose in mg/dL, mean (SD) | 105 (50) | 95 (34) | 93 (34) | 94 (28) | <0.001* |
| Total cholesterol in mg/dL, mean (SD) | 201 (46) | 193 (39) | 195 (39) | 197 (39) | <0.001* |
| *LDL in mg/dL, mean (SD)* | 123 (36) | 116 (33) | 118 (32) | 119 (33) | <0.001* |
| *HDL in mg/dL, mean (SD)* | 47 (13) | 45 (13) | 45 (13) | 46 (14) | 0.030* |
| *Triglycerides in mg/dL, mean (SD)* | 163 (165) | 166 (125) | 169 (152) | 167 (118) | <0.001* |
| Cardiovascular risk at 10 years in % |  |  |  |  |  |
| *Risk of CVD, P50 (P25 to P75)* | 8 (4 to 15) | 7 (4 to 15) | 7 (3 to 13) | 6 (3 to 12) | <0.001*** |
| *Risk of stroke or CHD, P50 (P25 to P75)* | 7 (3 to 13) | 5 (3 to 11) | 6 (3 to 10) | 5 (3 to 10) | <0.001*** |
| Notes: * ANOVA<br>** Chi2<br>*** Kruskal Wallis<br>Abbreviations:<br>CI, confidence interval; N/A, not applicable, CHD, coronary heart disease; CVD, cardiovascular disease; BMI, body mass index |  |  |  |  |  |

**Table 2.** Crude and adjusted associations between altitude and the risk of stroke and/or CHD at 10 years (linear regression model).

| Variable | Crude |  | Adjusted |  |  |  |
| --- | --- | --- | --- | --- | --- | --- |
| | $\beta$ (95% CI) | p-value | Saturated Model*<br>$\beta$ (95% CI) | p-value | Parsimonious Model**<br>$\beta$ (95% CI) | p-value |
| Constant | N/A | - | 14.22 (11.68 to 16.76) | <0.001 | 13.62 (12.62 to 14.67) | <0.001 |
| Altitude (per each extra meter) | -0.0005 (-0.0007 to -0.0003) | <0.001 | -0.0004 (-0.0006 to -0.0003) | <0.001 | -0.0005 (-0.0006 to -0.0003) | <0.001 |
| Female (male is the ref.) | -8.04 (-8.46 to -7.61) | <0.001 | -6.23 (-6.63 to -5.83) | <0.001 | -6.24 (-6.64 to -5.84) | <0.001 |
| Age (per each extra year) | 0.69 (0.65 to 0.74) | <0.001 | 0.60 (0.57 to 0.63) | <0.001 | 0.59 (0.56 to 0.62) | <0.001 |
| Ethnicity |  |  |  |  |  |  |
| Indigenous (ref.) | N/A | - | N/A | - | N/A | - |
| Afroecuadorians | 2.73 (1.32 to 4.14) | <0.001 | 0.93 (0.04 to 1.82) | 0.042 | 0.96 (0.07 to 1.85) | 0.035 |
| Montubio | 3.7 (2.16 to 5.20) | <0.001 | 0.52 (-0.44 to 1.47) | 0.287 | 0.51 (-0.44 to 1.46) | 0.295 |
| Mestizo | 2.90 (2.04 to 3.76) | <0.001 | 0.91 (0.35 to 1.47) | <0.001 | 0.94 (0.38 to 1.49) | 0.001 |
| Education level |  |  |  |  |  |  |
| Higher than secondary education (ref.) | N/A | - | N/A | - | N/A | - |
| Secondary education | -0.07 (-0.79 to 0.64) | 0.841 | -0.09 (-0.55 to 0.37) | 0.697 | - | - |
| Less than Primary education | 0.01 (-0.66 to 0.68) | 0.972 | -0.36 (-0.85 to 0.13) | 0.151 | - | - |
| Social security availability (not having is the ref.) | 1.24 (0.74 to 1.73) | <0.001 | -0.42 (-0.74 to -0.10) | 0.010 | -0.40 (-0.71 to -0.09) | 0.011 |
| Any immigrant in the family (no immigrant is the ref.) | -0.65 (-2.41 to 1.10) | 0.465 | 0.05 (-1.02 to 1.11) | 0.931 | - | - |
| Rural area (urban is the ref) | -0.99 (-1.48 to -0.50) | <0.001 | -0.22 (-0.58 to 0.14) | 0.225 | - | - |
| Income quintile |  |  |  |  |  |  |
| First quintile (the poorer group is the ref.) | N/A | - | N/A | - | N/A | - |
| Second quintile | 2.02 (1.27 to 2.77) | <0.001 | 0.83 (0.36 to 1.30) | 0.001 | 0.91 (0.45 to 1.38) | <0.001 |
| Third quintile | 2.38 (1.63 to 3.12) | <0.001 | 0.81 (0.31 to 1.31) | 0.002 | 0.99 (0.52 to 1.46) | <0.001 |
| Fourth quintile | 2.21 (1.47 to 2.96) | <0.001 | 0.65 (0.12 to 1.19) | 0.017 | 0.92 (0.44 to 1.40) | <0.001 |
| Fifth quintile (the richest) | 1.91 (1.16 to 2.65) | <0.001 | 0.93 (0.34 to 1.51) | 0.002 | 1.29 (0.80 to 1.77) | <0.001 |
| $\geq 7$ members in the family (otherwise is the ref.) | -0.38 (-1.02 to 0.25) | 0.238 | 0.01 (-0.39 to 0.40) | 0.971 | - | - |
| Any alcohol consumption | 3.36 (2.56 to 4.16) | <0.001 | 0.90 (0.40 to 1.40) | <0.001 | 0.89 (0.39 to 1.39) | <0.001 |
| (otherwise is the ref.) |  |  |  |  |  |  |
| History of hypertension | 2.64 (2.10 to 3.19) | <0.001 | 1.50 (1.15 to 1.84) | <0.001 | 1.50 (1.16 to 1.84) | <0.001 |
| BMI, in Kg/m<sup>2</sup> |  |  |  |  |  |  |
| ≥18.5 to <25 (ref.) | N/A | - | N/A | - | N/A | - |
| <18 | -2.30 (-5.66 to 1.06) | 0.181 | -1.17 (-3.21 to 0.87) | 0.262 | -1.11 (-3.15 to 0.93) | 0.287 |
| 25 to <30 | 1.75 (1.14 to 2.35) | <0.001 | 1.19 (0.81 to 1.56) | <0.001 | 1.19 (0.82 to 1.56) | <0.001 |
| ≥30 | 2.31 (1.66 to 2.96) | <0.001 | 1.97 (1.55 to 2.38) | <0.001 | 1.97 (1.56 to 2.38) | <0.001 |
| Hematocrit (per each extra % point) | 0.58 (0.53 to 0.63) | <0.001 | 0.14 (0.09 to 0.18) | <0.001 | 0.14 (0.09 to 0.18) | <0.001 |
| Triglycerides (per each extra unit in mg/dL) | 0.02 (0.02 to 0.02) | <0.001 | 0.02 (0.01 to 0.02) | <0.001 | 0.02 (0.01 to 0.02) | <0.001 |
| Notes:<br>* The value of the constant is the percentage of stroke risk and CHD at 10 years (according to Framingham equation) of a patient who lives in a mean altitude, is an indigenous person, not having social security, has higher than secondary education, who lives in an urban context, in who belongs to the first economic quintile, who has emigrant relatives, who lives with <7 persons, who does not drink alcohol, and did not reported high blood pressure as a medical diagnosis, has body mass index between 18.5 to <25 Kg/m<sup>2</sup>, has a mean hematocrit, and mean triglycerides concentration.<br>** The value of the constant is the percentage of risk of stroke and CHD at 10 years (according to Framingham equation) of a patient who lives in a mean altitude, is an indigenous person, has higher than secondary education, who belongs to the first economic quintile, who does not drink alcohol, and did not reported high blood pressure as a medical diagnosis, has body mass index between 18.5 to <25 Kg/m<sup>2</sup>, has a mean hematocrit, and mean triglycerides concentration.<br>Abbreviations:<br>CI, confidence interval; N/A, not applicable, CHD, coronary heart disease; CVD, cardiovascular disease; BMI, body mass index |  |  |  |  |  |  |

In the multivariable saturated model [adjusted for sex, age, ethnicity, educational level, availability of social security, immigrants in family, area, income quintile, overcrowding (≥ 7 inhabitants in the house), any alcohol consumption, history of hypertension, BMI, hematocrit, and triglycerides] participants had 0.0004 % less probability of developing cardiovascular (stroke and coronary heart disease) disease at 10 years *per* each increase in a meter in the altitude that participants live at (p<0.001) (**Table 2 and Figure 1**). This association persisted in the parsimonious model (β=-0.0005%, p=0.02). The effect of altitude on cardiovascular risk did not change after stratifying by sex (**S3 Table**).

The exclusion of people who were living at less than 53 m of altitude, those who were living at higher altitudes than 2635 m, people with self-reported diagnosis of hypertension, and those with elevated LDL cholesterol (≥160 mg/dL) in the model did not modify the estimate of the association between altitude and cardiovascular risk (**S4 Table**). Linear regression goodness of fit tests did not reveal any significant abnormality.

## Discussion

### Main findings

We found that, *per* each increase of 1000 m in the altitude that people live at, there is a reduction of almost half a percentual point in the cardiovascular risk at 10 years. This finding could be considered of little relevance from a clinical point of view, but altitude may be significant from a public health perspective, in terms of focusing with enhanced strategies of health promotion and primary prevention against CVDs in the population of coastal regions. In that regard, we share the criteria of Lopez-Pascual and cols. [18] about the potential existence of the Rose prevention paradox, where factors that make a small difference to the population distribution may be more important than factors that have a clinical impact on a small number of people [27].

### Contrast with scientific evidence

As several studies have demonstrated, altitude and specifically high altitude, is an important determinant of health outcomes, including cardiovascular disease. It is known that altitude-associated conditions may have different impacts on risk factors which can influence the development of different diseases; as well as having consequences on life expectancy and mortality [32]. For instance, a study conducted in the United States concluded that altitude may have protective effects on cardiovascular diseases and harmful effects on chronic pulmonary disease (COPD), with a dose-response relationship. Other authors [33], in a longitudinal, census-based record linkage study commented on the protective effect of living at higher altitude on coronary heart disease and stroke mortality; however, this study explained that this effect might have appeared due to climate-related factors and not necessarily due to classic cardiovascular risk factors. Another study in rural Greece concluded that living in mountainous areas have a possible protective effect from total and coronary mortality; this was explained by the possible effects of physical activity [34].

Even if a positive relation appears in some studies, a systematic review described that intermittent exposure to high altitude among workers increased the risk of CVDs. According to Burtscher, “COPD might be reduced when living at higher elevations but when suffering from COPD the mortality risk rises. Thus, COPD patients, especially when disease progresses would benefit from moving down to more oxygen rich sea level regions” [32]. As several circulatory changes occur during exposure to high altitude, some studies [19,35] concluded on the negative effect of altitude on populations suffering from pre-existing cardiovascular diseases.

It is not the first time that altitude has been linked with cardiovascular risk [17,25]. Some studies show that native Tibetans and Nepalese Sherpas rarely have systolic hypertension, having also lower levels of cholesterol [11]. A cohort study within the German-speaking part of Switzerland found a decreasing mortality by coronary heart disease or stroke when altitude increases. Some possible explanations to this finding are related to other risk factors such as pollution, since air pollution diminishes within increasing altitude. Another reason to support this fact is that diet could also be beneficial since food produced at higher altitudes might be richer in protective nutrients like omega 3 and vitamin D in dairy products [33].

Populations living in high lands experienced significant anatomic, physiological, and metabolic adaptation to cold temperature and low oxygen levels [13]. Other studies have suggested that living at high altitude may confer highlanders with protection against cardiovascular risk factors like dyslipidemia as well as CVDs such as hypertension and coronary ischemic disease [36,37]. The physiological changes are mostly due to ambient hypoxia, which occurs because of low atmospheric pressure, whilst the percentage of oxygen remains the same [14,38]. In Ecuador, there could also be an impact in lower cardiovascular risk, due to seasonal and latitudinal location. Studies have found that being close to the equator decreased blood pressure due to the correlation between Vitamin D absorption associated with UVB radiation [13].

Initial altitude-related adaptive changes include increased heart rate, cardiac output, hyperventilation, and, subsequently, increased red blood cell mass [14,19,39]. Eventually, a steady state could be reached, possibly due to the beneficial effects of ensuing cardiovascular, respiratory, and muscular adaptation mechanisms. However, it is important to note that there is substantial interindividual variability in the cardiovascular response to hypoxia [19]. In a study conducted by Sherpa et al, the female individuals living at high altitude had very low coronary heart disease risk, in addition to variations in the lipid profile of their population, namely a higher prevalence of hypertriglyceridemia in men [40]. Despite there is some evidence suggesting that there is a direct association between altitude and cardiometabolic risk [19,35,41,42], our study strongly supports the idea that altitude is a determinant of cardiovascular health in an inverse association when the exposure is prolonged.

Regarding the association between education and cardiovascular disease, we could not corroborate that link; and, although several studies have found that lower educational level is associated with higher cardiovascular risk, this relationship may vary depending on the analyzed cohort [43]. Furthermore, one possible explanation is that the association between education and CVD risk is mediated by obesity, metabolic syndrome, and high blood pressure [44]. Another explanation is that in Ecuador, the economic quintile could be a strong determinant of CVD risk, as it is seen in our analyses in which the poorest group had less CVD risk; thus, socioeconomic status is an important influence factor of CVD risk.

### Public health implications of our findings

Even though the clinical advice to live near the sea has become popular among cardiologists and other medical specialists when a patient suffers from cardiopulmonary diseases, our study gives valuable information that could guide and further analyse the need to reconsider or personalize this advice. Clearly, for patients with cardiovascular diseases who regularly live at low altitudes, there is sufficient evidence to not recommend trips to high altitudes, given the limited cardiopulmonary reserve which could become compromised and would probably increase their CVD risk; however, after our study, we can assume that individuals residing in elevated areas have a lower risk of developing CVDs. Consequently, considerations need to be made about exposing a patient to prolonged periods of time at high altitudes to receive the benefits of the stress produced to avoid cardiovascular disease.

Currently, it is not possible to make an appropriate recommendation for individual-based benefits for at-risk population of CVDs about exposure to high altitudes, but future studies are needed to explore whether simulated altitude conditions may have potential protective, or even, therapeutic interventions for cardiovascular risk [13]. It would be important to state that people who do not have a CVD can continue living at high altitude areas to enhance other health promotion and prevention measures against CVDs, contrary to the ones who have a CVD who could be advised to live at the sea level to improve pulmonary health problems.

Perhaps in the future, this evidence could be the door towards the development of new strategies for health promotion and primary prevention, possibly, also for treatments stratified to the altitude of the residential areas of different populations.

At the same time, the strengthening of the high-altitude food systems could be a way of promoting and spreading dietary practices of high-altitude regions. Mediterranean diet has been popularized among physicians, but it has serious limitations regarding its cost and availability in low- and middle-income countries like Ecuador. This study could be a useful element to think about promoting public health measures aimed to improve conditions for groups living in low-altitude areas. For example, to improve access to nutrients and educational policies directed to provide conditions to reduce the risk of CVDs, among others.

### Strengths and limitations

We are aware that this is a cross sectional study; therefore, it is not possible to establish a causal relationship between altitude and cardiovascular risk. At the same time, this is the first study, to our knowledge, which considered the impact of altitude on a meter-by-meter basis and its relation on formally calculated cardiovascular risk. In addition, this study considers the potential residual confounders of several factors from social demographic, economic, biochemical, and anthropometric measures, reducing the risk of confounding bias.

Regarding the possibility of residual confounding of physical activity (PA) in the association between altitude and CVDs risk, the amount of PA across altitude strata could be different, researchers have described significant adaptations to high altitude, in terms of body weight reduction, regardless of the amount of performed PA [45]; therefore, we do not believe that this is a significant source of bias. Even though we did not include PA in our analyses, the potential protective interaction between altitude and physical activity is an interesting hypothesis that goes far beyond this present study.

We could not separate the individuals and the coexistence of CVD, but when assessing the association excluding those with history of high blood pressure or highest values of cholesterol the association remained.

Other sources of potential residual confounding were the lack of information about pollution, humidity, temperature, and green space exposure. But, to our knowledge, most of those potential determinants of cardiovascular risk could not act as significant confounders as we found that the association was significant in all the sensitivity analyses.

Another limitation is related to the main quantitative nature of the study. Qualitative or mixed-methods tools could give an added value to the interpretation of the results as they provide further details of people’s lifestyles, behaviours, and contexts.

In addition, we did not consider the effects and/or interactions of place of birth and climate related determinants, as it was impossible to determine them within the available survey data. As a previous study indicated [33], being born at altitudes higher or lower than the place of residence was associated with lower or higher risk. Additionally, other studies elucidated on the effects of climate related factors such as temperature and levels of ultraviolet radiation [46] which have a possible negative effect on cardiovascular mortality. Consequently, including these factors in further research, could provide a better understanding of the risks, particularly important to the populations inhabiting the Andes.

Finally, information on food intake was not included in the survey questionnaire and information on occupation was not systematically collected. In the US population, the inverse association between altitude and obesity remained while adjusting for fruit and vegetable consumption [14].

### Suggestions for future research

It is necessary to elucidate on the epigenetic, molecular, and physiologic pathways to achieve protection against CVDs among people who live at higher altitudes. Regarding those protective mechanisms, several explanations have been proposed.

First, a study focused on assessing the effect of altitude training at 1600 and 1800 m showed larger improvements in performance after altitude training. It may be due to the greater overall load of training in hypoxia compared with normoxia, combined with a hypoxia-mediated increase in hemoglobin mass [47]. Nevertheless, there is uncertainty about the optimal timeframe for peak performances, suggesting that that the optimal window to improve physical performance is individual, and factors other than altitude exposure per se may be important.

Second, it is also possible that other factors such as polymorphisms for high altitude adaptation were involved in this effect, Valverde and cols. identified a number of single nucleotide polymorphisms (SNPs) exhibiting large allele frequency differences over 900,000 genotyped SNPs. A region in chromosome 10 (within the cytogenetic bands q22.3 and q23.1) was significantly differentiated between highland and lowland groups [48], interestingly, the results suggest that positive selection on the enhancer might increase the expression of this antioxidant, and thereby prevent oxidative damage. In addition, the most significant signal in a relative extended haplotype homozygosity analysis was localized to the SFTPD gene, which encodes a surfactant pulmonary-associated protein involved in normal respiration and innate host defence.

We believe that several potential issues to investigate would be relevant soon. There is a possibility that altitude induces expression of certain genes related to protective mechanisms against cardiovascular disease.

Moreover, we raised the question about how altitude could contribute to enhance the effects of population density-related features (pollution from urban areas, migrations or food accessibility) [18], several behavioural changes or treatments for primary prevention of cardiovascular risk and, even, for cardiovascular disease. In that regard, mortality may be profoundly modified by the complex interactions between behavioral and environmental conditions.

## Conclusion

From a public health perspective, altitude at which individuals live is an important health determinant of cardiovascular risk; specifically, *per* each increase of 1000 m in the altitude that people live at, there is a reduction of almost half a percentual point in the cardiovascular risk at 10 years. It is important to consider that living in the highlands could be a protective factor related to a reduced cardiovascular risk, but this effect could be not as relevant from a clinical perspective than from a public health point of view. Further research is necessary to understand genetics, epigenetic, and molecular mechanisms that explain this finding; complemented by further analyses on environmental, climate and behavioural factors.

## Data Availability

The Cardiovascular Risk Score project grants access to the main study results and data by making a request through the correspondence author.

## Acknowledgments

Authors thank the input from the “Cardiovascular Risk Score” project members. We, also, acknowledge the contributions from the *Instituto de Salud Pública* from PUCE.

## Author Contributions

Conceptualization: Andrea Cevallos Guerrero, Heidi Angela Fernández, Ángela León-Cáceres, Luciana Armijos-Acurio, and Iván Dueñas-Espín

Data curation: Carmenza Sevilla and Iván Dueñas-Espín

Formal analysis: Iván Dueñas-Espín

Funding acquisition: Luciana Armijos-Acurio, Carlos Erazo, Ruth Jimbo-Sotomayor, Hugo Pereira-Olmos, Henry Pineda-Abarca, Erika Quishpe-Narváez, Xavier Sánchez, Carmenza Sevilla, Betzabé Tello, Ana Torres-Castillo, Tatiana Villacrés, and Iván Dueñas-Espín.

Investigation: Andrea Cevallos Guerrero, Heidi Angela Fernández, Ángela León-Cáceres, Luciana Armijos-Acurio, Carlos Erazo, Ruth Jimbo-Sotomayor, Hugo Pereira-Olmos, Henry Pineda-Abarca, Erika Quishpe-Narváez, Xavier Sánchez, Carmenza Sevilla, Betzabé Tello, Ana Torres-Castillo, Tatiana Villacrés, and Iván Dueñas-Espín.

Methodology: Iván Dueñas-Espín.

Project administration: Iván Dueñas-Espín.

Writing – original draft: Andrea Cevallos Guerrero, Heidi Angela Fernández, Ángela León-Cáceres, and Iván Dueñas-Espín.

Writing – review & editing: Andrea Cevallos Guerrero, Heidi Angela Fernández, Ángela León-Cáceres, Luciana Armijos-Acurio, Carlos Erazo, Ruth Jimbo-Sotomayor, Hugo Pereira-Olmos, Henry Pineda-Abarca, Erika Quishpe-Narváez, Xavier Sánchez, Carmenza Sevilla, Betzabé Tello, Ana Torres-Castillo, Tatiana Villacrés, and Iván Dueñas-Espín.

## Supporting information

**S1 Fig. Flowchart diagram**.

**S2 Fig. Histogram of altitude**.

**S3. Fig. Histogram of Cardiovascular risk at ten years of stroke and coronary heart disease**.

**S1 Table. Characteristics of 3661 participants of Ecuadorian NHS included in the analyses**.

**S2 Table. Crude and adjusted Odds Ratios of high cardiovascular risk, calculated by four different equations, across altitude strata, according to multivariate logistic regression models**.

**S3 Table. Crude and adjusted associations between altitude and the risk of stroke and/or CHD at 10 years (linear regression model) stratified by sex**.

**S4 Table. Crude and adjusted associations between altitude and the risk of stroke and/or CHD at 10 years (linear regression models) excluding: (i) those who were living less than 53 m, (ii) those who were living higher than 2635 m, (iii) those with self-reported diagnosis of hypertension, and (iv) those with elevated LDL cholesterol (**≥**160 mg/dL)**.

