## Supporting information for "To what extent is the altitude at which we live associated with 10-year cardiovascular risk?"

*S1 Figure. Flowchart diagram.*

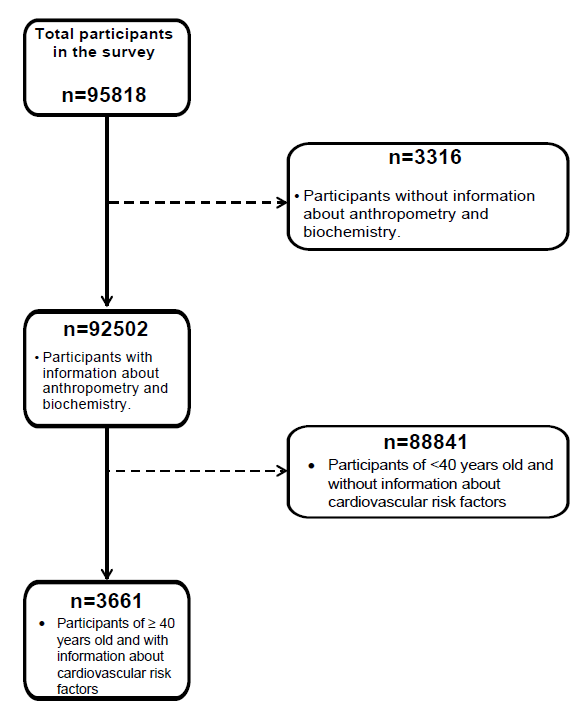

*S2 Figure. Histogram of altitude.*

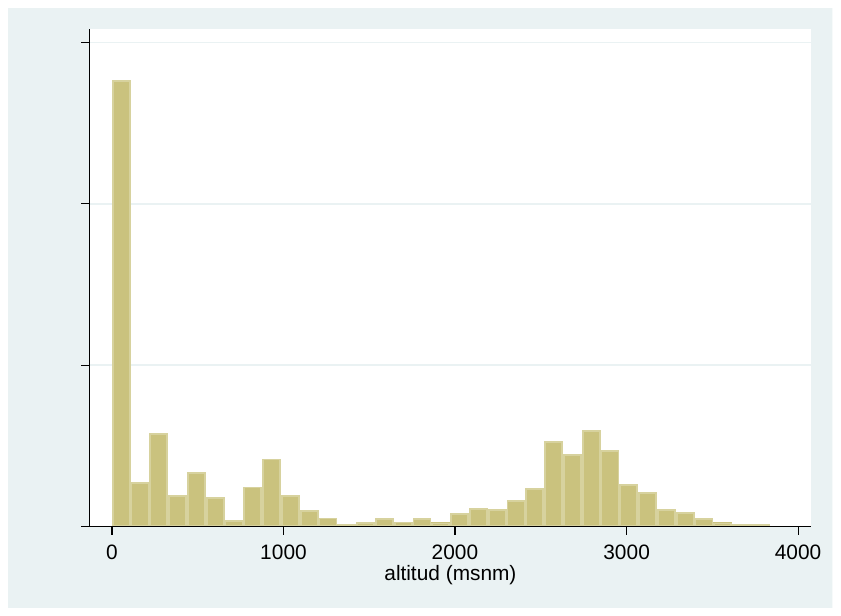

*S3 Figure. Histogram of Cardiovascular risk at ten years of stroke and coronary heart disease.*

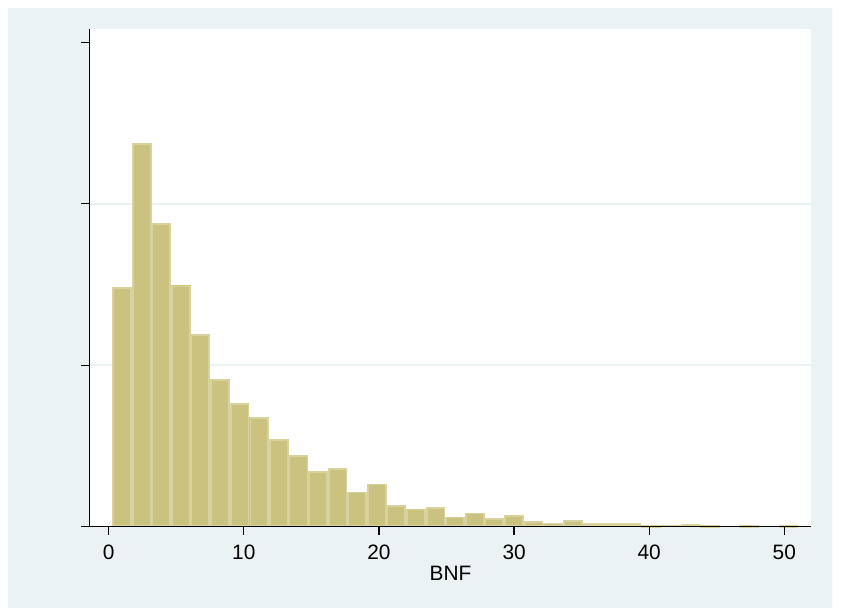

*S1 Table. Characteristics of 3661 participants of Ecuadorian NHS included in the analyses.*

| **Characteristics** | **All participants**  **n=3661** |
| --- | --- |
| Female sex, n (%) | 2532 (69) |
| Age, mean (SD) | 45 (7) |
| Altitude in m, P50 (P25 to P75) | 843 (53 to 2635) |
| Ethnicity |  |
| *Indigenous, n (%)* | 337 (9) |
| *Afroecuadorians, n (%)* | 160 (4) |
| *Montubio, n (%)* | 122 (3) |
| *Mestizo, n (%)* | 3042 (83) |
| Education level |  |
| *Higher than secondary education, n %* | 596 (16) |
| *Secondary education, n %* | 1244 (34) |
| *Primary or lesser education, n %* | 1821 (50) |
| Social security availability, n (%) | 1264 (35) |
| Any immigrant in the family (no immigrant is the ref.) | 67 (2) |
| Rural area (urban is the ref) | 1392 (38) |
| Income quintiles |  |
| *First quintile (the poorer group), n (%)* | 802 (22) |
| *Second quintile, n (%)* | 720 (20) |
| *Third quintile, n (%)* | 726 (20) |
| *Fourth quintile, n (%)* | 713 (19) |
| *Fifth quintile (the richest), n (%)* | 696 (19) |
| ≥ 7 members in the family, n (%) | 611 (17) |
| Harmful habits |  |
| *Alcohol consumption, n (%)* | 3267 (90) |
| *Tobacco smoking, n (%)* | 1643 (45) |
| Comorbidities |  |
| *History of hypertension, n (%)* | 858 (24) |
| *Diabetes as fasting glucose was ≥126 mg/dL, n (%)* | 180 (5) |
| Anthropometry |  |
| *Weight in Kg, mean (SD)* | 68 (13) |
| *Height in cm, mean (SD)* | 155 (9) |
| *BMI in Kg/m2, mean (SD)* | 28 (5) |
| *≥18.5 to <25, n (%)* | 772 (23) |
| *<18, n (%)* | 17 (<1) |
| *25 to <30, n (%)* | 1531 (45) |
| *≥ 30, n (%)* | 1050 (31) |
| Systolic pressure, mean (SD) | 122 (16) |
| Diastolic pressure, mean (SD) | 76 (10) |
| Laboratory Tests |  |
| *Hematocrit in %, mean (SD)* | 42 (5) |
| *Fasting glucose in mg/dL, mean (SD)* | 97 (38) |
| *Total cholesterol* in mg/dL*, mean (SD)* | 196 (41) |
| *LDL* in mg/dL*, mean (SD)* | 119 (34) |
| *HDL* in mg/dL*, mean (SD)* | 46 (13) |
| *Triglycerides* in mg/dL*, mean (SD)* | 166 (142) |
| Cardiovascular risk at 10 years in % |  |
| *Risk of CVD, P50 (P25 to P75)* | 7 (4 to 13) |
| *Risk of stroke and/or CHD, P50 (P25 to P75)* | 6 (3 to 11) |

*S2 Table.- Crude and adjusted Odds Ratios of high cardiovascular risk, calculated by four different equations, across altitude strata, according to multivariate logistic regression models.*

| **Altitude quartiles in m** | **Adjusted odds Ratios (95% CI) of cardiovascular risk*** | | | |
| --- | --- | --- | --- | --- |
|  | **≥20% of risk of stroke and CHD** | **p-value** | **≥20% of risk of CVD** | **p-value** |
| 0 to 53 (ref.) | 1 | - | 1 | - |
| 54 to 843 | 0.87 (0.54 to 1.43) | 0.59 | 0.88 (0.60 to 1.31) | 0.54 |
| 844 to 2635 | 0.58 (0.35 to 0.95) | 0.03 | 0.64 (0.43 to 0.95) | 0.02 |
| 2636 to 3834 | 0.43 (0.24 to 0.76) | <0.01 | 0.39 (0.24 to 0.62) | <0.01 |
| *p-for-trend* | 0.75 (0.63 to 0.90) | <0.01 | 0.75 (0.64 to 0.86) | <0.01 |
| Notes: *All models adjusted by ethnic group, education, economic quintile, any lifetime alcohol consumption, diagnosis of high blood pressure, body mass index, hematocrit, and triglycerides (see Table 2).  Abbreviations: CI, confidence interval; CHD = coronary heart disease. CVD=cardiovascular disease; m | | | | |

*S3 Table. Crude and adjusted associations between altitude and the risk of stroke and/or CHD at 10 years (linear regression model) stratified by sex.*

| **Variable** | **Only women**  **n=2532** | | **Only Men**  **n=** **1129** | |
| --- | --- | --- | --- | --- |
|  | **Parsimonious Model****  **β (95% CI)** | **p-value** | **Parsimonious Model****  **β (95% CI)** | **p-value** |
| Altitude (per each extra meter) | -0.0005 (-0.0007 to -0.0004) | <0.001 | -0.0003 (-0.0006 to <0.0001) | 0.075 |
| Age (per each extra year) | 0.49 (0.46 to 0.52) | <0.001 | 0.77 (0.71 to 0.83) | <0.001 |
| Ethnicity |  |  |  |  |
| *Indigenous (ref.)* | N/A | - | N/A | - |
| *Afroecuadorians* | 1.11 (0.21 to 2.01) | 0.016 | 1.33 (-0.56 to 3.22) | 0.168 |
| *Montubio* | 1.04 (<0.01 to 2.09) | 0.051 | 1.10 (-0.70 to 2.91) | 0.231 |
| *Mestizo* | 0.64 (0.08 to 1.19) | 0.024 | 1.91 (0.70 to 3.13) | 0.002 |
| Social security availability (not having is the ref.) | -0.42 (-0.73 to -0.10) | 0.010 | -0.53 (-1.15 to 0.10) | 0.098 |
| Income quintile |  |  |  |  |
| *First quintile (the poorer group is the ref.)* | N/A | - | N/A | - |
| *Second quintile* | 0.67 (0.21 to 1.14) | 0.005 | 1.33 (0.34 to 2.32) | 0.009 |
| *Third quintile* | 0.63 (0.16 to 1.10) | 0.009 | 1.49 (0.48 to 2.50) | 0.004 |
| *Fourth quintile* | 0.69 (0.22 to 1.16) | 0.004 | 1.04 (<0.01 to 2.09) | 0.005 |
| *Fifth quintile (the richest)* | 1.08 (0.60 to 1.56) | <0.001 | 1.59 (0.54 to 2.64) | 0.003 |
| Any alcohol consumption (otherwise is the ref.) | 0.71 (0.28 to 1.14) | 0.001 | 3.43 (1.41 to 5.46) | 0.001 |
| History of hypertension | 1.28 (0.94 to 1.62) | <0.001 | 2.07 (1.30 to 2.84) | <0.001 |
| BMI, in Kg/m^2^ |  |  |  |  |
| *≥18.5 to <25 (ref.)* | N/A | - | N/A | - |
| *<18* | -0.60 (2.68 to 1.48) | 0.571 | -1.54 (-5.74 to 2.66) | 0.471 |
| *25 to <30* | 0.44 (0.05 to 0.83) | 0.028 | 2.38 (1.63 to 3.12) | <0.001 |
| *≥ 30* | 0.91 (0.50 to 1.32) | <0.001 | 4.34 (3.40 to 5.29) | <0.001 |
| Hematocrit (per each extra % point) | 0.13 (0.08 to 0.17) | <0.001 | 0.19 (0.09 to 0.30) | <0.001 |
| Triglycerides (per each extra unit in mg/dL) | 0.02 (0.02 to 0.02) | <0.001 | 0.01 (0.01 to 0.02) | <0.001 |

*S4 Table. Crude and adjusted associations between altitude and the risk of stroke and/or CHD at 10 years (linear regression models) excluding: (i) those who were living less than 53 m, (ii) those who were living higher than 2635 m, (iii) those with self-reported diagnosis of hypertension, and (iv) those with elevated LDL cholesterol (≥160 mg/dL).*

| **Variable** | **Excluding those Living at < 53 m**  **n=2742** | | **Excluding those Living at ≥ 2635 m**  **n=2747** | | **Excluding those with self-reported diagnosis of hypertension**  **n=2803** | | **Excluding those with elevated LDL cholesterol (≥160 mg/dL)**  **n=3158** | |
| --- | --- | --- | --- | --- | --- | --- | --- | --- |
|  | **Parsimonious Model****  **β (95% CI)** | **p-value** | **Parsimonious Model****  **β (95% CI)** | **p-value** | **Parsimonious Model****  **β (95% CI)** | **p-value** | **Parsimonious Model****  **β (95% CI)** | **p-value** |
| Altitude (per each extra meter) | -0.0004 (-0.0005 to -0.0002) | <0.001 | -0.0005 (-0.0007 to -0.0003) | <0.001 | -0.0003 (-0.0004 to -0.0001) | <0.001 | -0.0004 (-0.0005 to -0.0002) | <0.001 |
| Female (male is the ref.) | -6.48 (-6.91 to -6.04) | <0.001 | -5.94 (-6.43 to -5.46) | <0.001 | -6.16 (-6.58 to -5.75) | <0.001 | -5.93 (-6.31 to -5.54) | <0.01 |
| Age (per each extra year) | 0.55 (0.52 to 0.59) | <0.001 | 0.62 (0.59 to 0.66) | <0.001 | 0.57 (0.54 to 0.60) | <0.001 | 0.55 (0.52 to 0.58) | <0.01 |
| Ethnicity |  |  |  |  |  |  |  |  |
| *Indigenous (ref.)* | N/A | - | N/A | - | N/A | - | N/A | - |
| *Afroecuadorians* | 0.35 (-0.77 to 1.47) | 0.541 | 1.05 (0.015 to 2.09) | 0.047 | 1.01 (0.03 to 1.99) | 0.044 | 0.70 (-0.12 to 1.56) | 0.110 |
| *Montubio* | 1.57 (0.40 to 2.73) | 0.008 | 0.60 (-0.47 to 1.68) | 0.270 | 0.60 (-0.42 to 1.63) | 0.250 | 0.06 (-0.84 to 0.96) | 0.899 |
| *Mestizo* | 0.93(0.39 to 1.47) | 0.001 | 1.14 (0.42 to 1.86) | 0.002 | 0.85 (0.29 to 1.41) | 0.003 | 0.79 (0.27 to 1.31) | 0.003 |
| Social security availability (not having is the ref.) | -0.30 (-0.64 to 0.03) | 0.075 | -0.31 (-0.68 to 0.06) | 0.102 | -0.27 (-0.59 to 0.04) | 0.088 | -0.33 (-0.62 to -0.03) | 0.031 |
| Income quintile |  |  |  |  |  |  |  |  |
| *First quintile (the poorer group is the ref.)* | N/A | - | N/A | - | N/A | - | N/A | - |
| *Second quintile* | 0.90 (0.41 to 1.39) | <0.001 | 0.91 (0.37 to 1.46) | 0.001 | 1.01 (0.53 to 1.48) | <0.001 | 0.63 (0.19 to 1.08) | 0.005 |
| *Third quintile* | 0.93 (0.42 to 1.44) | <0.001 | 0.92 (0.37 to 1.47) | 0.001 | 0.93 (0.44 to 1.42) | <0.001 | 0.88 (0.43 to 1.33) | <0.001 |
| *Fourth quintile* | 0.96 (0.43 to 1.49) | <0.001 | 0.79 (0.22 to 1.35) | 0.006 | 1.02 (0.53 to 1.51) | <0.001 | 0.66 (0.20 to 1.11) | 0.005 |
| *Fifth quintile (the richest)* | 1.22 (0.69 to 1.74) | <0.001 | 1.13 (0.53 to 1.73) | <0.001 | 1.23 (0.74 to 1.73) | <0.001 | 1.07 (0.60 to 1.53) | <0.001 |
| Any alcohol consumption (otherwise is the ref.) | 0.83 (0.30 to 1.36) | 0.002 | 0.90 (0.28 to 1.51) | 0.004 | 0.78 (0.27 to 1.29) | 0.003 | 0.92 (0.44 to 1.40) | <0.001 |
| History of hypertension | 1.41 (1.03 to 1.79) | <0.001 | 1.70 (1.29 to 2.11) | <0.001 | N/A | - | 1.36 (1.03 to 1.69) | <0.001 |
| BMI, in Kg/m^2^ |  |  |  |  |  |  |  |  |
| *≥18.5 to <25 (ref.)* | N/A | - | N/A | - | N/A | - | N/A | - |
| *<18* | -1.02 (-3.40 to 1.36) | 0.403 | -0.88 (-3.40 to 1.65) | 0.495 | -1.11 (-3.05 to 0.83) | 0.261 | -0.66 (-2.49 to 1.17) | 0.479 |
| *25 to <30* | 1.01 (0.61 to 1.42) | <0.001 | 1.23 (0.77 to 1.68) | <0.001 | 1.18 (0.81 to 1.55) | <0.001 | 1.06 (0.70 to 1.42) | <0.001 |
| *≥ 30* | 1.88 (1.43 to 2.34) | <0.001 | 2.03 (1.53 to 2.52) | <0.001 | 1.93 (1.50 to 2.35) | <0.001 | 1.72 (1.32 to 2.12) | <0.001 |
| Hematocrit (per each extra % point) | 0.12 (0.07 to 0.17) | <0.001 | 0.16 (0.10 to 0.21) | <0.001 | 0.08 (0.034 to 0.13) | 0.001 | 0.09 (0.05 to 0.13) | <0.001 |
| Triglycerides (per each extra unit in mg/dL) | 0.02 (0.0 2 to 0.02) | <0.001 | 0.01 (0.01 to 0.02) | <0.001 | 0.02 (0.01 to 0.02) | <0.001 | 0.02 (0.02 to 0.02) | <0.001 |
| Notes: * The value of the constant is the percentage of stroke risk and CHD at 10 years (according to Framingham equation) of a patient who lives in a mean altitude, is an indigenous person, not having social security, has higher than secondary education, who lives in an urban context, in who belongs to the first economic quintile, who has emigrant relatives, who lives with <7 persons, who does not drink alcohol, and did not reported high blood pressure as a medical diagnosis, has body mass index between 18.5 to <25 Kg/m^2^, has a mean hematocrit, and mean triglycerides concentration.  ** The value of the constant is the percentage of risk of stroke and CHD at 10 years (according to Framingham equation) of a patient who lives in a mean altitude, is an indigenous person, has higher than secondary education, who belongs to the first economic quintile, who does not drink alcohol, and did not reported high blood pressure as a medical diagnosis, has body mass index between 18.5 to <25 Kg/m^2^, has a mean hematocrit, and mean triglycerides concentration.  Abbreviations: CI, confidence interval; N/A, not applicable, CHD, coronary heart disease; CVD, cardiovascular disease; BMI, body mass index | | | | | | | | |
